# A Unique Clade of SARS-CoV-2 Viruses is Associated with Lower Viral Loads in Patient Upper Airways

**DOI:** 10.1101/2020.05.19.20107144

**Authors:** Ramon Lorenzo-Redondo, Hannah H. Nam, Scott C. Roberts, Lacy M. Simons, Lawrence J. Jennings, Chao Qi, Chad J. Achenbach, Alan R. Hauser, Michael G. Ison, Judd F. Hultquist, Egon A. Ozer

## Abstract

**Background:** The rapid spread of SARS-CoV-2, the causative agent of Coronavirus disease 2019 (COVID- 19), has been accompanied by the emergence of distinct viral clades, though their clinical significance remains unclear. Here, we aimed to investigate the phylogenetic characteristics of SARS-CoV-2 infections in Chicago, Illinois and assess their relationship to clinical parameters.

**Methods:** We performed whole-genome sequencing of SARS-CoV-2 isolates collected from COVID-19 patients in a Chicago healthcare system in mid-March, 2020. Using these and other publicly available sequences, we performed phylogenetic, phylogeographic, and phylodynamic analyses. Patient data was assessed for correlations between demographic or clinical characteristics and virologic features.

**Findings:** The 88 SARS-CoV-2 genome sequences in our study separated into three distinct phylogenetic clades. Clade 1 was most closely related to viral sequences from New York, and showed evidence of rapid expansion across the US, while Clade 3 was most closely related to those in Washington state. Clade 2 was localized primarily to the Chicago area with limited evidence of expansion elsewhere. At the time of diagnosis, patients infected with Clade 1 viruses had significantly higher average viral loads in their upper airways relative to patients infected with Clade 2 viruses, independent of time to symptom onset and disease severity.

**Interpretation:** These results show that multiple variants of SARS-CoV-2 are circulating in the Chicago area that differ in their relative viral loads in patient upper airways. These data suggest that differences in virus genotype impact viral load and may in turn influence viral transmission and spread.

**Funding:** Dixon Family Translational Research Award, Northwestern University Clinical and Translational Sciences Institute (NUCATS), National Institute of Allergy and Infectious Diseases (NIAID)

## INTRODUCTION

In late 2019, the first clusters of people with severe respiratory infections and pneumonia were reported in the Hubei province of China.^1-3^ The novel betacoronavirus known as severe acute respiratory syndrome coronavirus 2 (SARS-CoV-2) has since emerged as the cause of an unprecedented worldwide healthcare crisis with more than 7 million confirmed cases worldwide and more than 400,000 attributable deaths during the first six months of 2020.^4,5^ The spread of the novel coronavirus disease (COVID-19) has proceeded at different rates across the globe, but ongoing community transmission is widespread.

In the United States (US), the first confirmed case of COVID-19 was reported on January 20^th^ in Washington state with a second confirmed case reported shortly thereafter in Chicago, Illinois on January 23^rd^. Both cases were linked with recent travel to China.^6^ The Chicago patient later passed the virus to her husband in the first reported case of local transmission on January 30^th^.^6^ The state of New York reported its first case on March 1^st^ and rapidly developed into a hotspot. By the end of March, the US had confirmed cases in all 50 states and reported the most cases of any country in the world. Stay-at-home orders went into effect in an attempt to mitigate virus spread in Illinois, New York, and Washington on March 21^st^, 22^nd^, and 23^rd^, respectively.

Based on publicly available data, prevalence and mortality rates differ on a state-by-state basis [as of April 27th, 2020: New York, 282,143 cases (5.8% mortality); Washington state, 13,319 cases (5.5% mortality); Illinois, 41,777 cases (4.5% mortality)] (Supplemental Figure 1).^7,8^ These differences can be attributed to a multitude of factors including: geography, population density, demographics, the timing and extent of community mitigation measures, testing availability, healthcare infrastructure, and public health reporting practices.^9^ Whether variability in viral genotype may also contribute to these observed differences in transmission and disease burden remains unknown.

**Figure 1.**
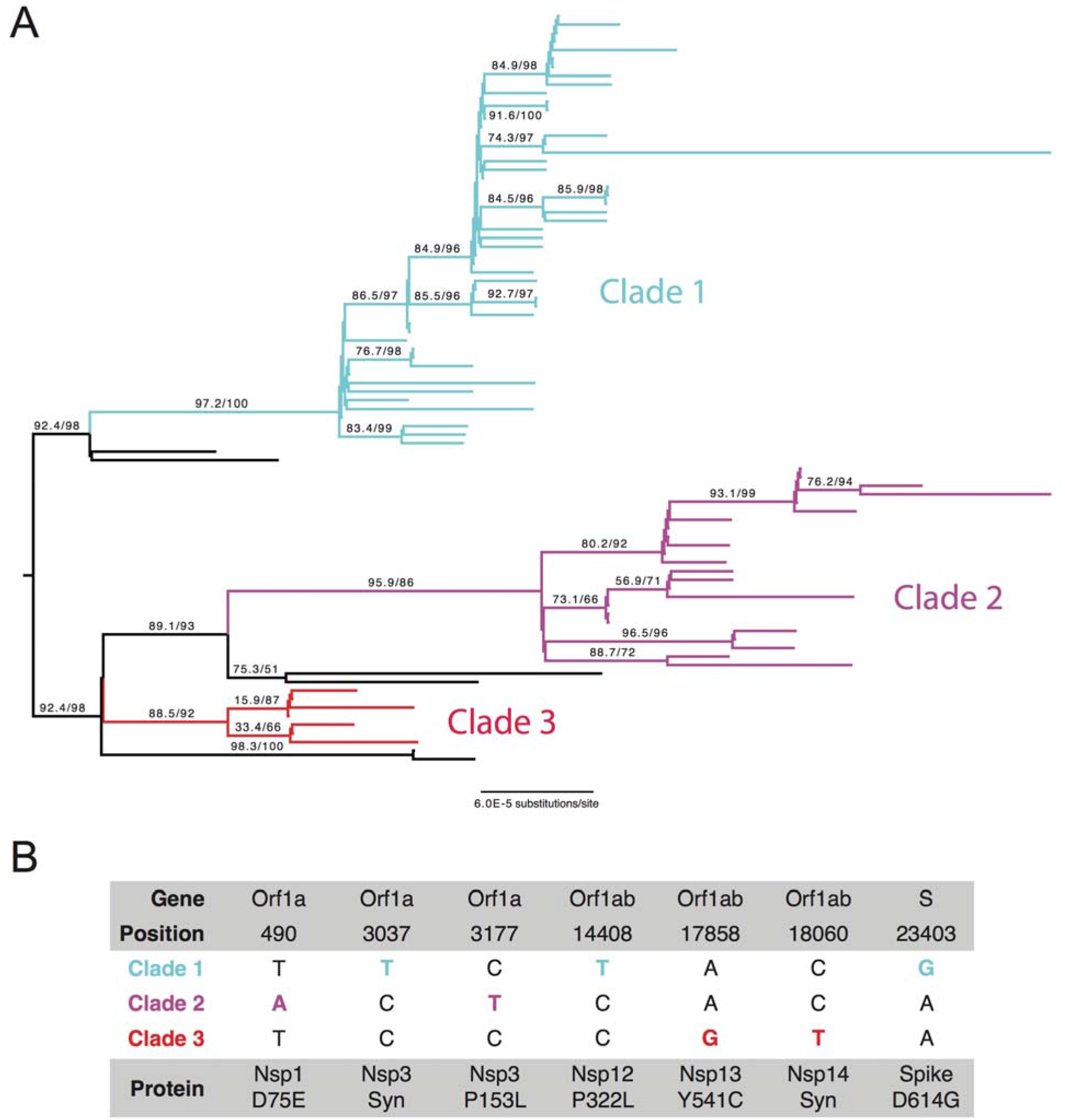
Phylogenetic Analysis of SARS-CoV-2 Isolates in Chicago. **a)** ML phylogenetic tree of 88 SARS-CoV-2 specimen genomes from Northwestern Memorial Hospital and Lake Forest Hospital. All non-zero statistical support values for each branch are indicated. Black branches represent sequenced isolates that do not belong to any of the three major clades. **b)** Clade- defining mutations at the US level. Positions are numbered according to the reference genome (NC_045512.2).

In parallel with the unprecedented speed and scale of the pandemic, the sharing and integration of clinical and epidemiological data has also occurred at an exceptional pace, with over 40,000 viral genome sequences of SARS-CoV-2 from across the globe currently available via online databases such as GISAID.^10^ Rapid whole-genome sequencing (WGS) of emerging viruses allows for informed development of molecular diagnostic assays, as well as tracing of patterns of spread across multiple epidemiological scales.^11^ Understanding the molecular epidemiology of the virus through evolutionary models and phylogenetic analysis can further aid in estimating the genetic variability and evolutionary rate, which can have significant implications for disease progression as well as drug and vaccine development.

While extensive WGS has been reported in Washington state and New York, there are relatively few sequences available from Illinois. Here, we present a phylogenetic analysis of 88 SARS-CoV-2 viruses from the airways of COVID-19 patients seeking care in a single healthcare system in Chicago, Illinois from March 14, 2020 through March 21, 2020. We further assessed these viral genomic data for correlations with clinical and demographic features of our cohort.

## METHODS

### Specimen collection and testing

Individuals suspected of COVID-19 within two hospitals of the Northwestern Medicine healthcare system were screened for SARS-CoV-2 infection as per the CDC protocol.^12^ Samples were collected between March 14, 2020 through March 21, 2020. Participants included individuals presenting to emergency departments at Northwestern Memorial Hospital (NMH) or Lake Forest Hospital (LFH), patients admitted to the hospital at NMH, and outpatient or corporate health clinic patients. Nasopharyngeal swab and/or bronchoalveolar lavage fluid specimens were collected from screened individuals and stored in either phosphate-buffered saline (PBS) or viral transport medium. Viral RNA was extracted from clinical specimens utilizing the QIAamp Viral RNA Minikit (Qiagen). Clinical testing for SARS-CoV-2 presence was performed by quantitative reverse transcription and PCR (qRT-PCR) with the CDC 2019-nCoV RT-PCR Diagnostic Panel utilizing N1 and N2 probes as previously described.^12^ All specimens with a cycle threshold (Ct) less than or equal to 35 were considered positive and included in this study. Ct values from the N1 probes were used in all subsequent analyses.

### Whole Genome Sequencing and analysis

Detailed whole genome sequencing and analysis methodologies can be found in the Supplemental Methods. Briefly, cDNA synthesis was performed with SuperScript IV First Strand Synthesis Kit (Thermo) according to manufacturer’s specifications. Direct amplification of the viral genome cDNA was performed using multiplexed primer pools per protocols provided by the Artic Network (version 3 release) ^13^. The sequencing library preparation protocol was adapted from the ARTIC Network protocol and Oxford Nanopore protocol “PCR tiling of COVID-19 virus”.^13-15^ Adapter ligation and cleanup was performed using the Oxford Nanopore Genomic DNA by Ligation kit according to manufacturer specifications. Libraries were sequenced on the Nanopore MinION device using FLO-MIN106D Type R9.4.1 flow cells.

Base calling of nanopore reads was performed on the Northwestern University’s Quest High Performance Computing Cluster for Genomics using Guppy v3.4.5 with the DNA R9.4.1 450bps High Accuracy configuration and default quality score filtering (--qscore_filtering). Read quality filtering, reference alignment, and consensus sequence generation was performed using ARTIC Network software and protocols.^15,16^ Regions with less than 10-fold read coverage were masked. We chose to use a lower read coverage cutoff than the default of 20 in the ARTIC Network software to allow for consensus sequence calls in regions with relatively poor amplification due to inefficient primer binding. Manual validation of variant calls in regions with 10 – 19x coverage was performed using Tablet software.^17^ Consensus genome sequences were deposited in the GISAID database (accession numbers provided in Supplemental Table 1).

### Phylogenetic and statistical analysis

Detailed phylogenetic and statistical methodologies can be found in the Supplemental Methods.

### Ethics Statement

This study was reviewed and approved by the Institutional Review Board of Northwestern University.

### Role of the Funding Sources

The funding sources had no role in the design of the study, in the collection/analysis/ interpretation of the data, in the writing of the report, or in the decision to publish. The corresponding authors had access to all of the data in the study and had final responsibility for the decision to submit this work for publication.

## RESULTS

### Collection characteristics and sequencing results

Between March 14 and March 21, 2020, a total of 127 qRT-PCR clinical tests performed at NMH were positive for SARS-CoV-2 (Supplemental Figure 2A). Residual RNA from these diagnostic tests was collected for WGS. Of these, 37 specimens failed to achieve sufficient amplification required for sequencing or had inadequate read coverage after sequencing. Almost half of the 90 successfully sequenced specimens had been collected from patients evaluated in emergency departments at NMH or LFH, 25% from non-ICU hospitalized patients at NMH, and 12% from patients in the NMH ICU (Supplemental Figure 2B). Two sequenced specimens were found to be redundant and were excluded from further analysis, leaving 88 sequenced specimens from 88 individuals. The qRT-PCR cycle threshold (Ct) values of these specimens ranged from 14.52 to 32.37. After applying alignment quality filters and normalizing segment coverage to a maximum of 200 reads in each direction, the average median genome coverage across non-overlapping amplicon sequences was 348 reads per specimen (min 56, max 395).

**Figure 2.**
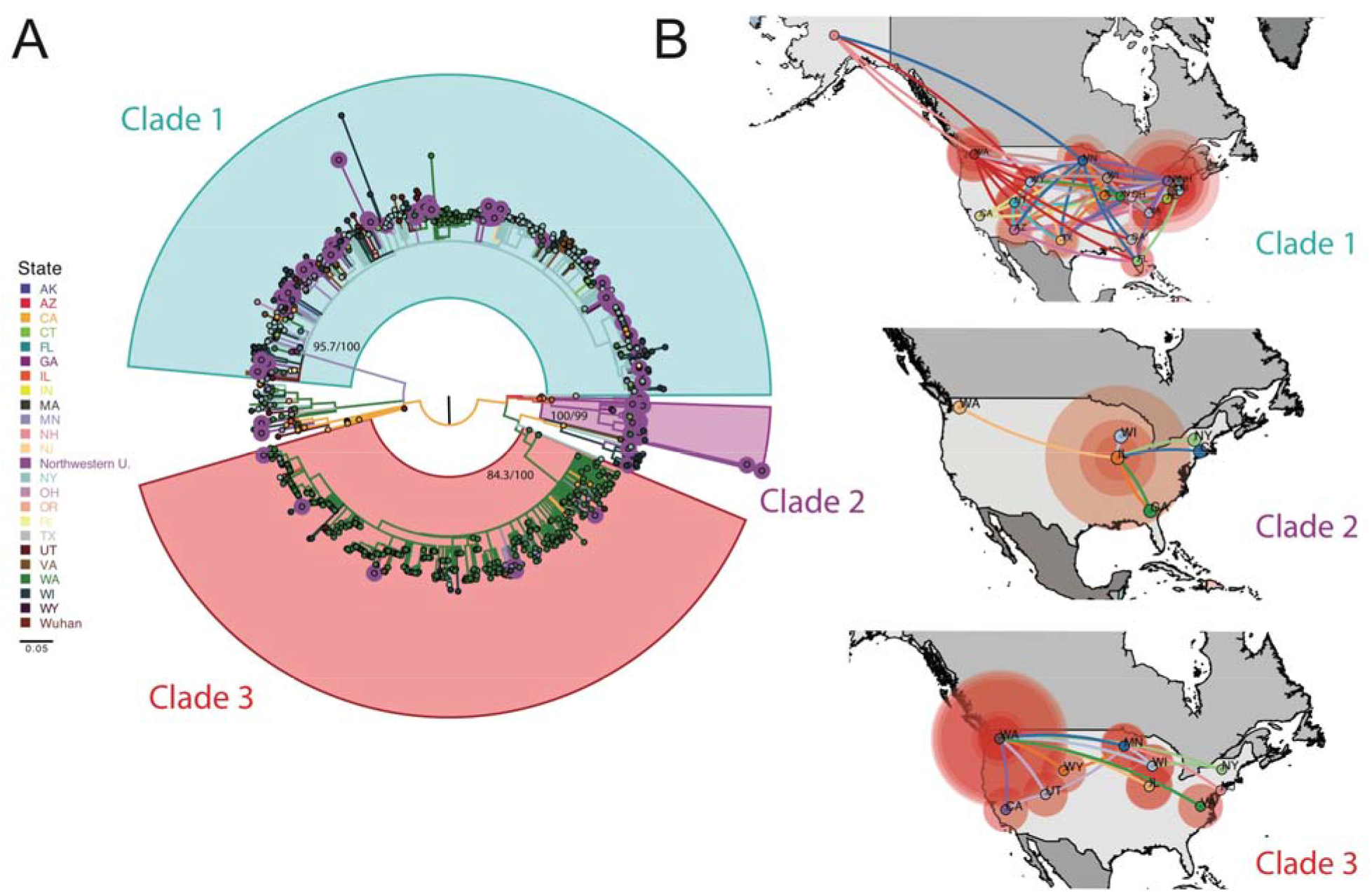
Phylogenetic and Phylogeographic Analysis of Chicago Isolates compared to the US Epidemic. **a)** ML phylogenetic reconstruction of full genome sequences from the United States. We included sequences from Northwestern and sequences from US available in GISAID. Branches are colored by location and tips corresponding to Northwestern sequences are highlighted. Well-supported clades of the tree that include our defined Chicago clades are indicated. **b)** Phylogeographic patterns of US isolates in three major clades represented in the Chicago collection under a discrete diffusion model. Westward movements are indicated by lines with an upward curvature, eastward movements are indicated by lines with a downward curvature, lines are colored according to the most probable geographical location of their descendent node, and circle sizes around a node are proportional to the number of lineages maintaining that location.

### Three main clades are detected through phylogenetic inference

We first performed a maximum likelihood (ML) phylogenetic analysis of the 88 SARS-CoV-2 specimen genomes in this study (Supplemental Methods). This analysis showed that most of the sequences (93%) clustered in three main clades (support: >85% aLRT and >85% UFboot) (Figure 1A). For the purposes of this report we will refer to these clades, ordered by relative abundance in the sequence set, as Clades 1 (n=51, 57.9%), 2 (n=24, 27.3%), and 3 (n=7, 7.9%). Using the Pangolin software method of lineage classification,^18,19^ isolates in Clade 1 were assigned mainly to lineage B·1 (92% assigned to lineage B·1; 2% to B·1.1; and 6% to B·1.5), the majority of isolates in Clade 2 belonged to lineage A·3 (92% to A.3; 8% to A), and isolates in clade 3 belonged mainly to lineage A·1 (86% to A·1; 14% to A) (Supplemental Table 1). The intra- and inter-clade variability of the sequences showed a mean intra-group variability of 1.16×10^−4^ base differences per site [SEM= 3.16×10^−5^] versus a mean inter-group variability of 4.80 ×10^−4^ base differences per site [SEM= 1.17×10^−4^], confirming the higher within-clade sequence similarity in the observed phylogenetic clustering.

### Clade 2 viruses are uniquely prominent in the Chicago area

To place the three observed clades in a broader geographic context, we expanded our analysis to include 901 SARS-CoV-2 sequences from the US deposited in GISAID as of April 4, 2020. With the complete US dataset, we performed ML phylogenetic reconstruction of the sequences, generated a temporal phylogeny, and reconstructed the ancestral states of the tree to identify clade-defining mutations (Figure 1B) and possible geographical transitions. These analyses confirmed our previous clustering, with a bulk of the Chicago sequences falling into three distinct clades (Figure 2A). Overall, the local epidemic in the Chicago area was broadly reflective of the epidemic in the US as a whole. Clade 1 viruses clustered predominantly with those from New York, while Clade 3 viruses clustered with those circulating on the West Coast, particularly in Washington state. These two clades showed a broader spread across the US than Clade 2, which was comprised almost exclusively of our newly sequenced specimens in Illinois with only a few representatives from elsewhere in the US (Figure 2A, Supplemental Figure 3). Clade 2 also contained a few previously sequenced SARS-CoV-2 specimens identified in Illinois at the end of January 2020.^6^

**Figure 3.**
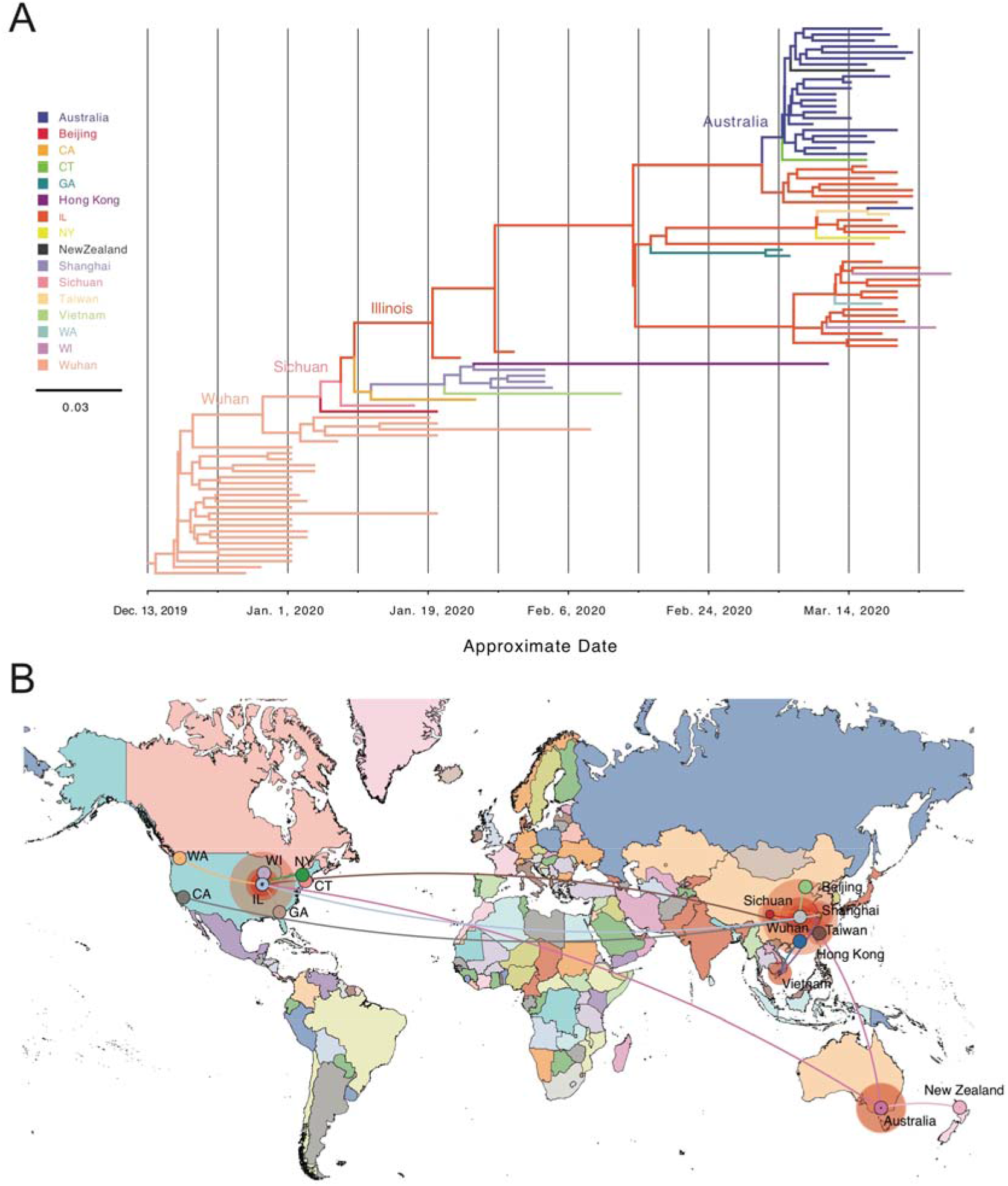
Phylogenetic and Phylogeographic Analysis of Chicago Isolates compared to the Global Pandemic. **a)** Phylodynamic tree of US and global Clade 2 related genome sequences. We used global sequences phylogenetically related to Clade 2 available in the GISAID database and performed the analysis encompassing simultaneous estimation of sequence and discrete (geographic) trait data. The depicted phylogenetic tree corresponds to the maximum clade credibility tree. Branch colors represent the most probable geographical location of their descendent node inferred through Bayesian reconstruction of the ancestral state. The width of the branches represents their posterior probability. X-axis corresponds to the inferred date. **b)** Phylogeographic reconstruction of the origin of Clade 2 under a discrete diffusion model. Westward movements are indicated by lines with an upward curvature, eastward movements are indicated by lines with a downward curvature, lines are colored according to the most probable geographical location of their descendent node, and circle sizes around a node are proportional to the number of lineages maintaining that location.

We next studied the phylodynamic and phylogenomic characteristics of these three clades using a Bayesian approach to infer possible differences in their population dynamics. These analyses suggested very limited spread of Clade 2, with an average proportion of branches residing in Illinois of 0.81 [95% CI: 0.66-0.88] (Supplemental Figure 3B). Illinois was found to be the most probable origin of Clade 2 in the US with an estimated most recent common ancestor (TMRCA) around January 18, 2020 [95% Highest Posterior Density (HPD) interval: January 9 - January 21, 2020]. The timing, location, and the sequences close to the inferred root of this tree (USA_IL_1 and USA_IL_2) coincide with the first reported COVID-19 cases in Illinois,^6^ though alternate early introductions cannot be ruled out. These results suggest that Clade 2 was introduced to the Chicago area early in the US epidemic with limited spread beyond Illinois (Figure 2B, middle panel).

A similar analysis of Clade 1 suggested multiple introductions, a high degree of diversity, and the existence of several probable sub-clades. Although the highest proportion of branches were found to reside in New York, this proportion was relatively low at 0.34 [95% CI: 0.31-0.37], in agreement with the broad geographic spread of this clade (Supplemental Figure 3A). These findings are most consistent with Clade 1 being centered in New York, but rapidly expanding throughout the region and to many other locations across the US (Figure 2B, bottom panel). The TMRCA of this clade in the US was calculated to be February 10, 2020 [95% HPD interval: January 28 - February 20, 2020], an estimated date in close alignment with prior reports.^20^ These results are suggestive of this clade having had multiple introductions into the Chicago area with some occurring early during the epidemic in February and March of 2020.

Finally, US isolates in Clade 3 are estimated to have originated in Washington state and remained centered there throughout the investigated time course (proportion of branches residing in Washington of 0.83 [95% CI: 0.81-0.85]) (Supplemental Figure 3C). This clade was also estimated to have been introduced into the US early in the epidemic (TMRCA = January 16, 2020 [95% HPD interval January 11 - January 17, 2020]), though we cannot rule out the possibility that this clade was independently introduced on more than one occasion.^21^ These data are most consistent with subsequent expansion of Clade 3 from Washington state to multiple locations throughout the US, including Illinois, followed by limited spread elsewhere (Figure 2B, top panel).

Overall, our results suggest that as of mid-March 2020, three main clades of SARS-CoV-2 were circulating in the Chicago area. Clade 2 appears to have been introduced in Illinois early in the epidemic and seems to have undergone more limited spread when compared to Clade 1 centered in New York and Clade 3 centered in Washington.

### Clade 2 has limited global distribution

To further investigate the origin and population dynamics of the Clade 2, we performed a phylogenetic analysis at the global level. We included an additional 3099 SARS-CoV-2 sequences deposited in GISAID from outside the US up through April 4, 2020. We conducted similar ML analyses as those performed on the US sequences above (Supplemental Figure 4). These results further confirmed strong statistical support for Clade 2 (91.6% aLRT and 92% UFboot). Again, the detected spread of this clade has been exceptionally limited, with a majority of sequences derived from this study, China, or Australia. Ancestral reconstruction of these global sequences showed low divergence of these samples from those first sequenced in Wuhan, China (Figure 3A).

**Figure 4.**
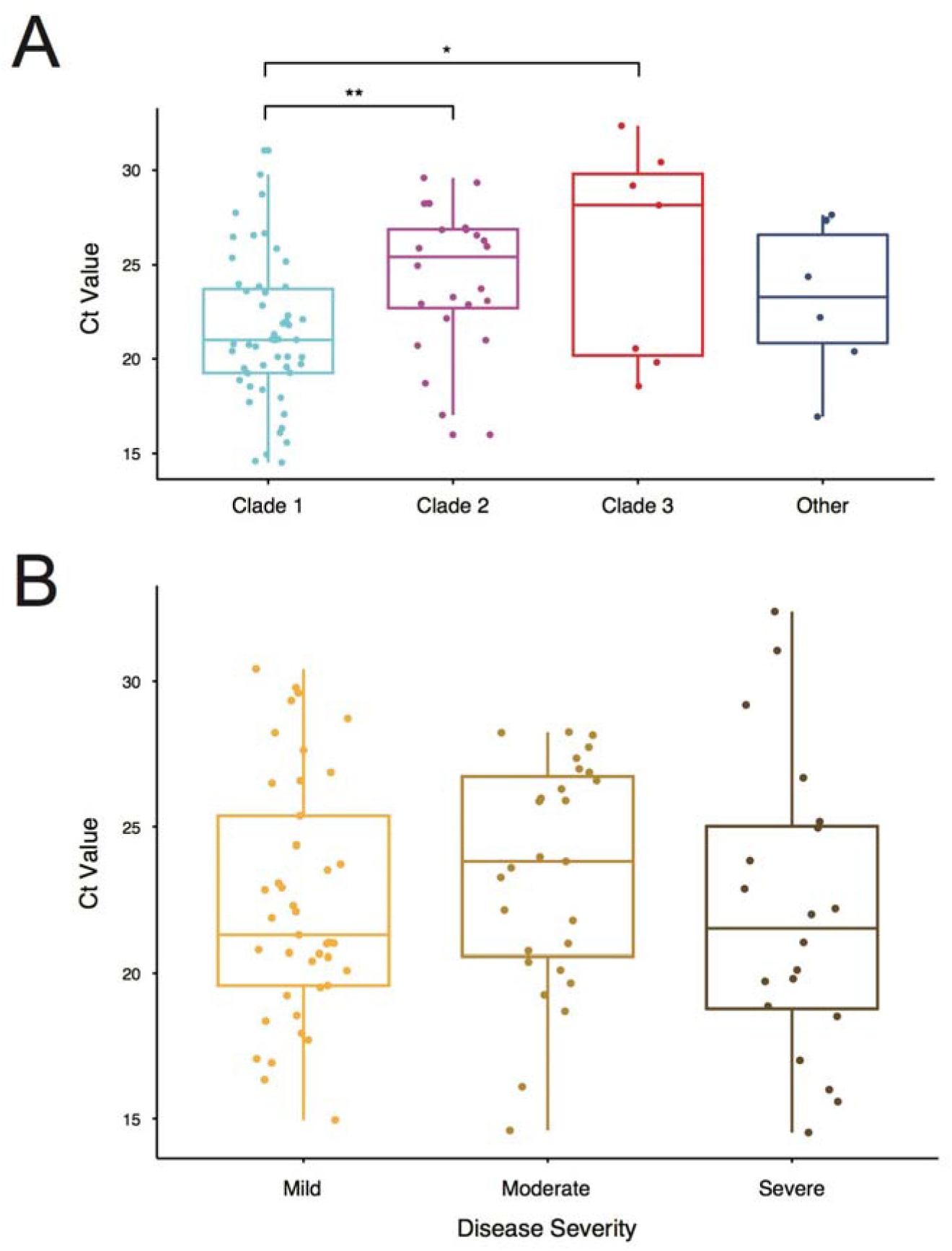
Associations between Viral Clade and Ct Value and Disease Severity. **a)** PCR Cycle threshold (Ct) values of patient samples grouped by major Clade assignment. **b)** Specimen Ct values by maximum disease severity. Mild (blue) = no hospital admission; Moderate (red) = hospital admission, but no ICU stay; Severe (green) = ICU admission. In both panels, horizontal lines in each box represent the median value and the lower and upper error bars are the interquartile ranges. Significance is indicated for the comparisons performed within each fitted model (* = q-value<0.05; ** = q-value <0.01).

To better discern the most likely origin of Clade 2, we performed a phylogeographical analysis using all available Clade 2 sequences and related global sequences. As suggested above, these results determined the most likely origin of this clade to be China at our current state of global sampling (Figure 3B). Clade 2-like sequences from other areas outside the US most likely represent introductions of similar viruses into those areas rather than secondary seeding from Illinois. One exception to this is a cluster of sequences in Australia that appear to be more closely related to those in Illinois than China (Figure 3B). Together, these findings are most consistent with an introduction of Clade 2 into Illinois from China in January 2020. Additionally, global analysis with the other two clades (Supplemental Figure 4) suggest that the New York- centered Clade 1 was likely to have been introduced into the US from Europe while the Washington-centered Clade 3 was most likely introduced directly from China, consistent with other reports.^21^

Together, these analyses show distinct global phylodynamics of the three main clades observed in the Chicago area: Clade 1 was introduced by way of New York through Europe, Clade 2 was introduced directly from China, and Clade 3 was introduced through Washington state from China. While Clade 1 is well-represented on the East Coast and Clade 3 is well-represented on the West Coast, to date Clade 2 is almost exclusively found in the Chicago area.

### Clade 2 viruses are associated with lower viral titers at the time of diagnosis

The limited spread of Clade 2 SARS-CoV-2 viruses both in the US and worldwide could suggest phenotypic variability that may impact transmission rate. To further investigate this, we compared the Ct values for each sample, which serves as a proxy for viral load in the patients’ upper airways at the time of diagnosis. We grouped the viruses according to their assigned clade, creating an additional group for the viruses that did not fall into any of the three major clades. We fit a linear model to test for differences in Ct values between the clades, controlling for the date of isolation, hospital location, age, sex, race, severity of illness, and specimen collection source (Supplemental Table 2). We detected significantly higher Ct values corresponding to lower viral loads in Clade 2 (Chicago-centered) compared to Clade 1 (New York-centered, q=0.008) and Clade 3 (Washington-centered, q=0.027) (Figure 4A). Due to the limited sample sizes for Clade 3 (n = 7) and ungrouped specimens (n = 6), our subsequent analyses focus solely on Clades 1 (n = 51) and 2 (n = 24).

Next, we tested whether the source of the sample (nasopharyngeal swab [NP] vs bronchoalveolar lavage fluid [BAL]) influenced the viral load, as measured by the Ct value, and/or could account for the differences between Clades 1 and 2. We tested for differences between sample source, clade, and both, accounting for their potential interaction in the linear model. We found a significant difference between NP and BAL samples (q= 0.0484), with lower Ct values in the BAL specimens (Supplemental Figure 5A). When considering only NP samples, the Ct values of Clade 1 viruses were still significantly lower than Clade 2 viruses as observed previously (q=0.0062). However, no significant difference in Ct value was detected by clade in the BAL samples. While the numbers of BAL samples are low, this result suggests that the viral load differences between the two clades are primarily observed in the upper airways of infected patients.

It has been reported that viral loads in the upper airways generally decrease over the course of infection.^22,23^ To determine whether the difference in average Ct values between samples from patients infected with Clade 1 or Clade 2 viruses corresponded with the time of specimen collection, the number of days between reported symptom onset and specimen collection was determined for each patient from available case files. No significant difference in time from symptom onset to specimen collection between patients infected with Clade 1 or Clade 2 viruses was observed (Supplemental Figure 5B). Additionally, the date of possible virus exposure was able to be determined for 29 patients and again no significant difference was found between clades (Supplemental Figure 5C). Consistent with prior reports, we did find that later dates of specimen collection after symptom onset significantly correlated with higher Ct values among patients infected with Clade 1 viruses, though this same trend was not observed among patients infected with Clade 2 viruses (Supplemental Figure 5D). These results suggest that the average viral load in the upper airways of patients infected with Clade 2 viruses is lower relative to those infected with Clade 1 viruses at the time of diagnosis independent of time since symptom onset.

Given early reports that COVID-19 disease severity is correlated with SARS-CoV-2 viral load,^24,25^ we next looked for associations between viral Clades 1 and 2 and maximal disease severity. Disease severity was binned as mild (outpatient or ED only), moderate (inpatient hospitalization), or severe (ICU admission and/or death) and set as the outcome variable. We then fit a logistic regression model including all available demographic and clinical variables. Interestingly, we found no significant association between any available variable and disease severity in our dataset, including clade (Figure 4B). We finally fit a logistic regression model including all available demographic and clinical variables with clade as the outcome variable (Supplemental Table 3). Consistent with our previous results, Ct value was the only variable associated with viral clade and vice versa. These data suggest that Ct value in the upper airway at the time of diagnosis may not be associated with disease severity, though it is associated with the viral clade.

## DISCUSSION

In this study, we report the phylogenetic and phylodynamic analyses of 88 SARS-CoV-2 genomes from COVID-19 patients in Chicago, Illinois, US, which largely fall into three major clades. Two of these three clades are broadly representative of the major circulating clades that expanded and disseminated rapidly across the US through early April 2020 with points of origin in New York (Clade 1) and Washington state (Clade 3). In addition, we identified a third major clade (Clade 2) that is relatively unique in the US and predominantly found in the Chicago area based on current sampling. Viruses in this clade lack the rapid and wide dispersion evidenced with the other viral clades. Notably, patients infected with Clade 1 viruses presented with significantly higher viral loads in the upper airways at the time of diagnosis compared to patients infected with Clade 2 viruses. These data suggest that differences in SARS-CoV-2 genotype may impact viral load, which in turn may influence transmission and overall viral spread.

Chicago, Illinois is the third-largest city in the US and is located centrally in the country with direct flights to all continents except Antarctica. Despite recording the second confirmed case of COVID-19 and the first confirmed case of person-to-person transmission in the US, relatively little SARS-CoV-2 sequencing data has been reported from the Chicago area with only 11 sequences available on GISAID [as of May 11, 2020]. The data presented here add an additional 88 sequences to this collection and constitute the first systematic analysis of SARS-CoV-2 whole genome sequences from COVID-19 patients in the Chicago area.

Phylogenetic analyses of these sequences demonstrate that the major clades circulating in Chicago in mid-March 2020 represent separate introduction events that can be traced back to distinct sources. Although the Chicago-centered Clade 2 and the Washington-centered Clade 3 are phylogenetically related, this analysis shows that they were the result of geographically and temporally separate introductions into the US from China. Our analysis suggests that Clade 2 viruses were introduced into the Chicago area from China and began community transmission around mid-January 2020. While Clade 3 viruses are related, they are more consistent with an independent introduction event through an intermediate in Washington state. We additionally find that that Clade 1 viruses are most likely to have been introduced to Chicago via New York, through Europe, a model which is also supported by a recent pre-print report describing the European origin of SARS-CoV-2 in New York City.^20^

Both the US and global sequence data suggest relatively little dispersion of Clade 2 strains beyond Illinois as of mid- to late-March 2020. Indeed, though the clade appears to have originated from China as early as mid-January 2020, there is little evidence to date of spread beyond a few likely cases in other Midwest states and a secondary cluster in Australia. This is in contrast to Clade 3 and especially Clade 1 viruses, which have spread over a larger geographic area in less time. Though a large number of isolates collected in the US and globally were available to this study through the GISAID collective, under-sampling of some regions and/or patient groups could have resulted in imprecise estimates of transmission patterns or potentially have missed a broader distribution of Clade 2 specimens. However, the proportion of viruses sampled from Chicago in this study that were Clade 2 (24 of 88, 27.3%) was far greater than the proportion in the database sampled across the US (10 of 901, 1.1%) or from non-US countries (34 of 3099, 1.1%), suggesting that Clade 2 was more abundant in the Chicago area relative to other sampled locations as of late March 2020. As more genomes both from Illinois and elsewhere are sequenced, we will continue to assess for changes in the population structure and distribution of SARS-CoV-2.

Notably, we found that Clade 2 viruses had significantly lower viral loads in patient upper airways relative to Clade 1 viruses at the time of diagnosis. This is among the first reports of SARS-CoV-2 phylogenetic lineage correlating with phenotypic differences. Clade 1 viruses are defined by nonsynonymous mutations in the RNA-dependent RNA polymerase (Nsp12 P322L) and Spike (S D614G) proteins. Other studies have likewise linked the Spike D614G mutation to a broader global spread and to higher viral loads.^26,27^ Similarly, other reports have noted the rapid spread of the Nsp12 P322L mutation.^28^ Our study was not designed to determine causal relationships between mutations and phenotypes, and it is possible that the virus is adapting multiple, independent changes that aid in its propagation. Lower viral loads in the upper airways of infected patients may decrease viral transmissibility and impact the overall spread, but further laboratory and clinical studies will be required to test the impact of these variants on viral replication, disease severity, and transmissibility.

Other than the potential for lower viral loads, there are other factors that could account for the muted spread of Clade 2 viruses outside the Chicago region. First, the state of Illinois instituted a relatively rapid and stringent lockdown by implementing restrictions on mass gatherings on March 13^th^, closing schools on March 17^th^, and issuing stay-at-home orders on March 21^st^.

While these restrictions went into place concurrent with sample gathering for this study, they may have contributed to a slower spread of Clade 2 viruses outside the state. Second, the virus may be more prevalent among members of a relatively more insular community within the state with comparatively less travel or socialization with non-community members. Third, these viruses may be found to have distinct clinical manifestations that lessen the opportunity for asymptomatic transmission to multiple other individuals. Ongoing studies of clinical and demographic patient data associated with ongoing specimen collection may provide support for these or other scenarios.

The other clinical and demographic data we examined, including disease severity, did not correlate significantly with clade or Ct value. Previous reports suggested that disease severity may be correlated with viral load as measured by Ct value during disease, but our data suggest the same may not hold true for Ct values collected at diagnosis.^29^ Although patients in the ICU had lower Ct values than patients in other locations, supporting a link between viral load and severity, most ICU samples were BAL specimens, not NP specimens. This correlation may therefore simply be reflective of different Ct values in the lower vs. upper airways. Finally, we did observe a correlation between Ct value and the time since symptom onset, confirming previous reports,^22,23^ though this did not change significantly by clade and did not explain the higher Ct values of Clade 2 viruses. Further studies with more specimens and associated clinical data will be needed to elucidate these relationships.

As the specimens in this study were collected over the first 8 days during which testing was broadly available at our institution, the phylogenetic patterns and abundances observed represent a cross-sectional snapshot of our region. Longitudinal evaluations could reveal more detailed clade dynamics and population structure. Additionally, these specimens were collected from one healthcare system primarily serving patients from the North and West sides of Chicago, potentially limiting generalizability to other regions in and around Chicago or other parts of Illinois. Nevertheless, these data suggest Chicago is an ideal location for observational and interventional studies to capture the breadth of SARS-CoV-2 genetic diversity. This ability to compare diverse viral genotypes in a single hospital setting allowed us to identify clade-specific differences in viral load at the time of diagnosis, which may in turn have consequences for transmissibility.

## Data Availability

Genome sequence data that support the findings of this study have been deposited in GISAID (https://www.gisaid.org/) with the accession codes given in Supplemental Table 1

## ACKNOWLEDGEMENTS

This research was supported in part through the computational resources and staff contributions provided for the Quest high performance computing facility at Northwestern University, which is jointly supported by the Office of the Provost, the Office for Research, and Northwestern University Information Technology. Funding for this work was provided by: a Dixon Translational Research Grant made possible by the generous support of the Dixon family (E.A.O. and J.F.H.); a COVID-19 Supplemental Research award from the Northwestern Center for Advanced Technologies (NUCATS - M.G.I., C.J.A., and J.F.H.), the Gilead Sciences Research Scholars Program in HIV (J.F.H.), and NIH grants K22 AI136691 (J.F.H.), U19 AI135964 (E.A.O., A.R.H.), K24 AI04831 (A.R.H.), and T32 AI095207 (H.H.N., S.C.R.).

## AUTHOR CONTRIBUTIONS

Conceptualization: R.L.R., H.H.N., S.C.R., L.M.S., A.R.H., M.G.I., J.F.H., E.A.O. Investigation: R.L.R., H.H.N., S.C.R., L.M.S., J.F.H., E.A.O. Resources: H.H.N., S.C.R., C.Q., L.J.J., A.R.H., M.G.I., J.F.H., E.A.O. Formal analysis: R.L.R., H.H.N., S.C.R., L.M.S., E.A.O. Supervision: L.J.J., A.R.H., M.G.I., J.F.H., E.A.O. Funding acquisition: A.R.H., M.G.I., J.F.H., E.A.O. Writing: R.L.R., H.H.N., S.C.R., L.M.S., A.R.H., M.G.I., J.F.H., E.A.O.

## DECLARATION OF INTERESTS

The authors declare no competing financial interests.

## Notes

### Competing Interest Statement

The authors have declared no competing interest.

### Author Declarations

This study was reviewed and approved by the Institutional Review Board of Northwestern University.

